# Societal Uses of the Main Water Bodies Inhabited by Malaria Vectors and Implications for Larval Source Management

**DOI:** 10.1101/2024.05.29.24308146

**Authors:** Najat F. Kahamba, Felista Tarimo, Khamisi Kifungo, Winifrida Mponzi, Siaba A. Kinunda, Alfred Simfukwe, Salum Mapua, Betwel Msugupakulya, Francesco Baldini, Heather M. Ferguson, Fredros O. Okumu, Marceline F. Finda

## Abstract

**Introduction:** Larval source management (LSM) can effectively suppress mosquito populations at source and provides an opportunity to address major challenges such as insecticide resistance that undermine primary interventions like insecticide-treated nets (ITNs). While mostly implemented in urban and arid settings, emerging research indicates its potential in some rural settings in east and southern Africa, where the main malaria vector, *Anopheles funestus*, prefers permanent and semi-permanent water bodies that support year-round transmission. Targeting these unique habitats could amplify effectiveness of LSM but requires careful considerations of local societal practices and expectations - particularly since mosquito breeding sites often also serve as community water resources. The aim of this study was therefore to explore how the societal uses of aquatic habitats by local communities in rural south-eastern Tanzania might influence LSM strategies, focusing on habitats frequented by *An. funestus*.

**Methods:** This study was conducted in three villages in the Ulanga and Malinyi districts of southeastern Tanzania using a mixed-methods approach. Quantitative data were collected through a cross-sectional surveillance of all aquatic habitats, while qualitative data were gathered via a combination of individual unstructured interviews, focus group discussions with various community groups and field observations of community practices and activities. Data analysis employed weaving and inferencing techniques to integrate findings from both quantitative and qualitative components, thereby developing a comprehensive understanding from the respondents’ perspectives.

**Results:** A survey of 931 aquatic habitats revealed that 73% contained mosquito larvae, with late instar An. funestus identified in 23% of these habitats. River streams segments were the most common habitat type, accounting for 41%, followed by ground pools at 4%; other types included pits, rice fields, ditches, and puddles. Community use was noted for 90% of these habitats, including 95% of those with An. funestus larvae, for activities such as domestic chores such as cooking, washing utensils, washing clothes and bathing, agriculture, livestock rearing, brickmaking, and fishing. Focus group discussions indicated community readiness to implement LSM, favoring larviciding and habitat manipulation over habitat removal. Community concerns regarding LSM centered on the safety of larvicides for animal and human health and their environmental impact. The discussions proved the need for LSM interventions to integrate seamlessly with daily activities; and for community education on LSM safety and efficacy.

**Conclusion:** This study offers valuable insights into community perspectives on LSM for malaria control in rural settings, emphasizing the dual role of aquatic habitats as both mosquito breeding sites and community water sources. This presents a set of unique challenges and opportunities – suggesting that LSM strategies must address both the biological aspects of mosquito control and the socio-economic realities of local communities. Notably, there was a marked preference for larviciding and habitat manipulation over habitat removal, with a strong emphasis on health and environmental safety. Overall, the study highlights the critical importance of educating communities, adopting culturally sensitive approaches to LSM, and aligning LSM strategies with the needs, perspectives, and daily lives of local communities.

## Introduction

Over the past two decades, significant progress has been made in the fight against malaria, primarily due large-scale deployment of preventative and therapeutic measures (Bhatt *et al*., 2015; Hemingway *et al*., 2016; World Health Organisation, 2021). Vector control strategies, notably insecticide-treated bed nets (ITNs) and indoor residual spraying (IRS), have been at the forefront; accounting for over 70% of the progress achieved (Bhatt *et al*., 2015). Despite these advancements, malaria remains a public health concern in sub-Saharan Africa, with some areas seeing unchanged or increasing case numbers (World Health Organisation, 2021). Among other challenges, malaria control efforts are being complicated by the rise of drug-resistant parasites, the spread of insecticide resistance in mosquitoes, and mosquito behavior adaptations that reduce the effectiveness of existing controls (Russell *et al*., 2011; Govella and Ferguson, 2012; Matowo *et al*., 2017).

In response to these ongoing challenges, the World Health Organization (WHO) recommends, among other strategies, larval source management (LSM) as a supplementary intervention in malaria-endemic countries across Africa (World Health Organization, 2022). This approach is increasingly recognized for its potential in the malaria control arsenal, though there are still multiple uncertainties and conflicting statements about its viability (Fillinger and Lindsay, 2011).

*Anopheles funestus*, one of the most efficient malaria vectors, has contributed significantly to the persistence of malaria due to its adaptability and widespread presence (Coetzee and Fontenille, 2004; Lwetoijera *et al*., 2014; Msugupakulya *et al*., 2023). Understanding the mosquito life cycle is crucial for appreciating the relevance of LSM. The mosquito life cycle includes four stages: egg, larva, pupa, and adult, with the first three stages being aquatic (Rejmánková *et al*., 2013; Jacques Derek Charlwood, 2020).

LSM disrupts the mosquito lifecycle through three primary approaches: i.) habitat modification, which involves the complete removal of breeding sites, for example, filling the breeding habitat with sand or constructing structures to eliminate it entirely; ii.) habitat manipulation, involving routine activities to make environments less conducive to mosquito breeding, for example flushing streams, removing vegetation and debris, and exposing habitats to the sun; and iii.) larviciding, the application of biological or chemical insecticides to water to halt larval development (Organization, 2013).

By targeting the mosquito populations at its source, LSM can be particularly relevant for overcoming challenges such as insecticide resistance that diminish the efficacy of conventional vector control measures like insecticide-treated nets (ITNs). Additionally, the strategic use of LSM offers a way to manage mosquito populations effectively, without solely relying on chemical interventions (Fillinger and Lindsay, 2011). Indeed, microbial larvicides, like *Bacillus thuringiensis israelensis (bti)* and *Bacillus sphaericus (bs),* have been effective and can overcome problems like insecticide resistance and environmental damage often associated with other chemical treatments (Kandyata *et al*., 2012; Tusting *et al*., 2013). At its core, the approach reduces mosquito populations and, as a result, can effectively suppress malaria transmission (Fillinger and Lindsay, 2011; Tusting *et al*., 2013; Derua *et al*., 2019), and reduced incidence of malaria (Fillinger *et al*., 2008; Tusting *et al*., 2013).

While LSM holds significant promise in the fight against malaria, its adoption by global funding bodies has encountered several obstacles. For example, the World Health Organization (WHO) recommends LSM for areas where suitable mosquito breeding sites are few, fixed and findable (FFF) (Hemingway *et al*., 2016; World Health Organisation, 2021). Because of these guidelines, LSM is currently mostly implemented in urban and arid settings. However, in many malaria-endemic regions, these larval habitats are abundant, widespread, and often located in areas that are difficult to access, making the implementation of LSM strategies difficult. Additionally, larviciding, one of the key components of LSM, is often costly and labor-intensive (Dambach *et al*., 2019; Berlin Rubin *et al*., 2020). Another challenge facing larviciding is the diversity nature of malaria vectors and their unique aquatic habitat usage, making it difficult to address all vectors simultaneously and effectively with this approach (Nambunga *et al*., 2020; Kahamba *et al*., 2024).

In southeastern Tanzania and other regions of the country, *An. funestus* has emerged as a major vector in malaria transmission, accounting for about 90% of the overall entomological inoculation rate (EIR) (Kaindoa *et al*., 2017; Matowo *et al*., 2021). This trend is also seen across other parts of east and southern Africa, where the species contributes majority of ongoing transmission (Msugupakulya *et al*., 2023). Given the unique traits of *An. funestus*, such as its breeding in fixed permanent and semi-water bodies, which persist into dry months and can help sustain year-round malaria transmission (Kahamba geospatial paper), LSM is argued to be a potential strategy against this vector. On account of this unique ecological suitability, and the fact that the adults, despite being highly resistant to insecticides, remain mostly endophilic and endophagic, a combined approach of LSM and adulticides such as dual-active ITNs or non-pyrethroid IRS, has been suggested as particularly valuable for not only reducing *An. funestus*-mediated malaria transmission but potentially even crushing the local populations this this species (Kahamba *et al*., 2022).

Targeting these unique habitats could be significantly magnify the impact of LSM in these rural settings, but it requires an indepth understanding of the interactions between communities in malaria-endemic areas and the aquatic habitats of malaria vectors. Insights into how communities use these habitats, and their overall opinions can shape the way larval source management (LSM) strategies are designed and implemented (Lupenza *et al*., 2021). For example, if communities regularly use the same habitats for drinking, bathing, or other daily activities, they may be strongly against habitat removal but supporting larviciding, especially if they report a biting nuisance from these habitats and have information on the safety of the approach.

Many studies from different locations have demonstrated the correlation between community engagement and LSM success (van den Berg *et al*., 2018; Berlin Rubin *et al*., 2020; Gowelo, 2020; Lupenza *et al*., 2021; Hakizimana *et al*., 2022; Gowelo *et al*., 2023), indicating the need for strategies that are adapted to meet community experiences and needs. Efforts should therefore be made to ensure that LSM practices adequately account for local societal experiences, needs and expectations, especially since the same water bodies where mosquitoes breed tend to be the same as those used by communities for other purposes.

The aim of this study was therefore to explore how the use of aquatic habitats by local communities in rural south-eastern Tanzania might influences LSM strategies, focusing on habitats frequented by *An. funestus*. To achieve this, we first identified and quantified the main aquatic habitats used by local malaria vector species, and assessed if and how these habitats were being used by local communities. Lastly, we assessed community perspectives and recommendations on LSM approaches for malaria vector control.

## Methods

### Study area

This study was conducted in three malaria endemic communities in sout-eastern Tanzania, namely Ikungua and Chikuti villages in Ulanga district, and Sofi Majiji village in Malinyi district (Fig. 1). Detailed description of these communities is provided by Kahamba *et al* 2024 (Kahamba et al., 2024). The residents primarily engage in subsistence farming and pastoralism, with small groups involved in artisanal mining, brick making, fishing, and small-scale business like food vendors, general stores, and market. Rice cultivation occurs year-round, depending on natural rainfall during the rainy season and irrigation during the dry season (Peel, Finlayson and McMahon, 2007). Other food crops include maize, beans, sesame seeds, and cassava. These villages are situated at an altitude of approximately 300-450 m above sea level, with major rivers like the Ruli river providing essential water sources for irrigation and daily use. Access to electricity and clean water is limited, so most residents depend on shallow wells for domestic water needs. Cooking is mostly done using wood or charcoal. The environmental features, including the presence of major rivers and low-altitude settings, contribute to mosquito ecology by providing abundant breeding sites, thus influencing malaria transmission dynamics in these areas.

**Figure 1:**
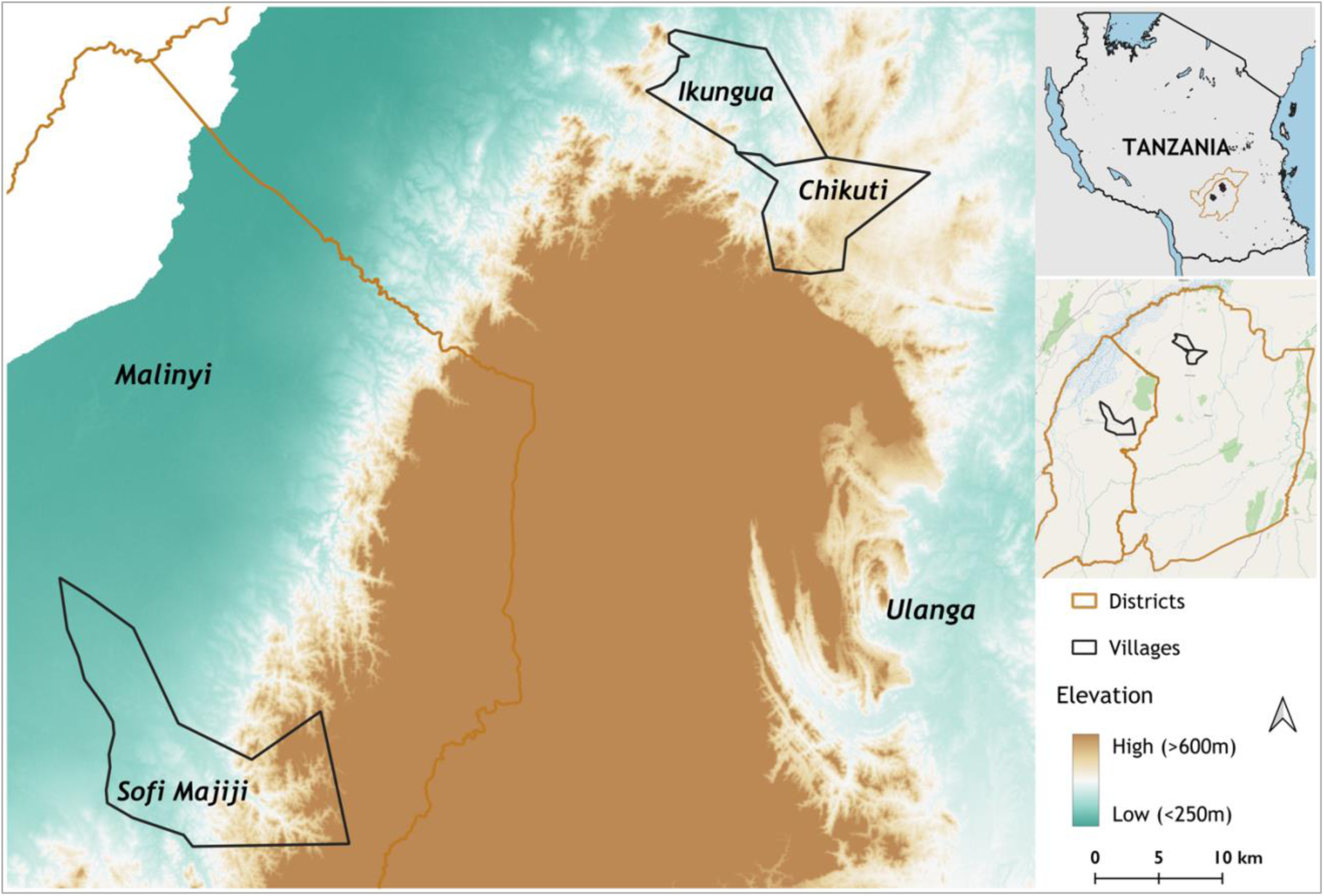
A map showing study villages in Ulanga and Malinyi districts.

**Figure 2:**
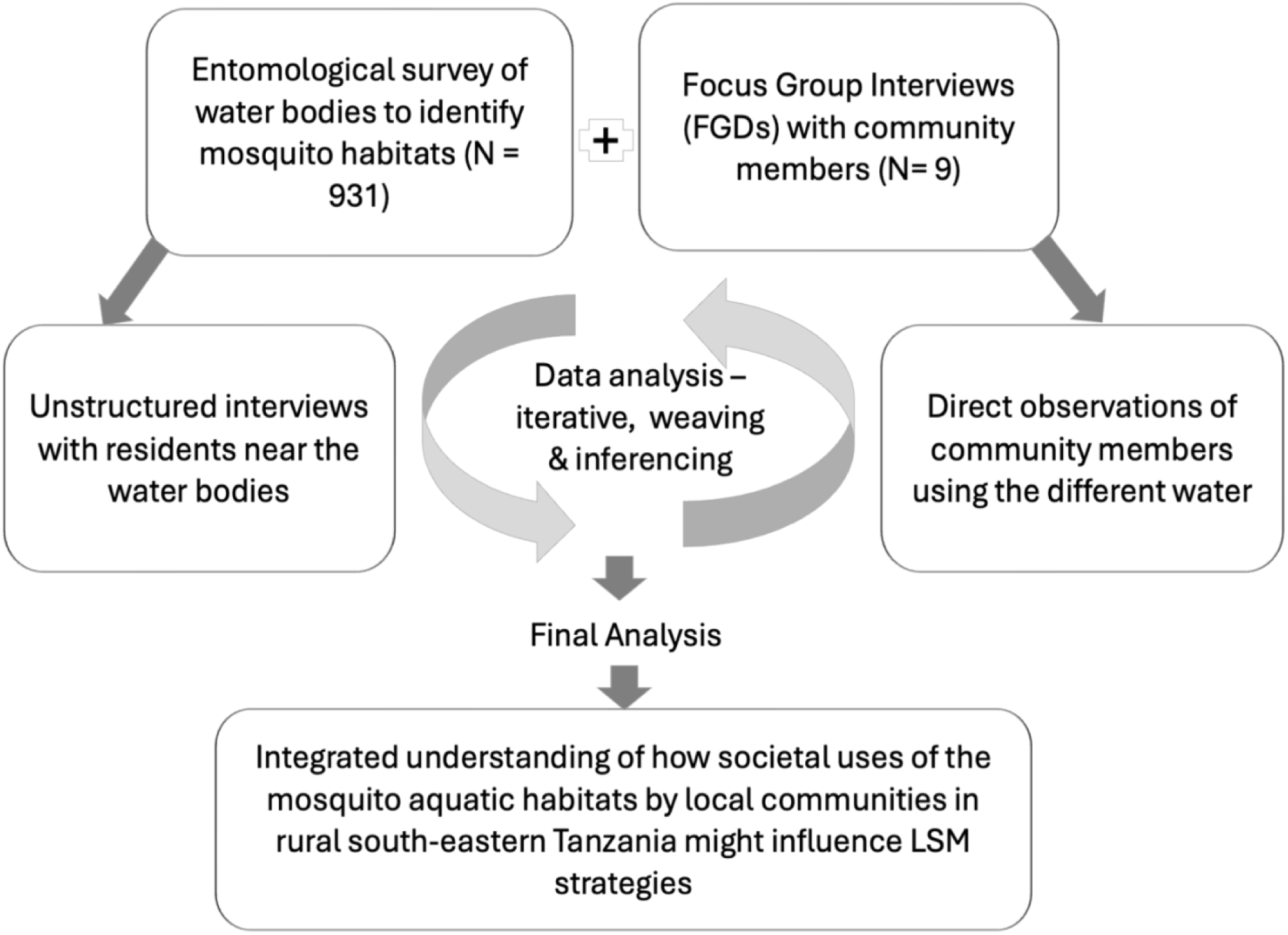
Mixed-Methods Approaches for data collection and analysis of how the societal uses of aquatic habitats by local communities in rural south-eastern Tanzania might influence LSM strategies, focusing on habitats frequented by *An. funestus*

### Study design

This study used a sequential mixed-method research design for the main study objectives. Quantitative data was collected first during the dry season ( July - November 2022), which included assessment of aquatic habitats, respective densities of *An. funestus* larvae or pupae, and domestic uses for the aquatic habitats. Subsequently, the qualitative component included a series of focus group discussions (FGDs) to explore communities’ perspectives and recommendations on how the LSM approaches can be integrated to their daily practices was conducted in November 2022.

Additionally, field visits and direct observations were made to assess the actual community practices and uses of these habitats.

### Habitat characterization and entomological surveys

Quantitative data collection followed the procedure detailed in Kahamba *et al* 2024 (Kahamba *et al*., 2024). A cross-sectional larval survey was done to identify and characterize aquatic habitats containing *An. funestus.* This process involved recording various environmental characteristics such as habitats type, size, watercolour, permanence of water, water movement, water source, presence of shades, presence of vegetation, and the presence of algae (Fig. 3). Immature mosquitoes were collected using either 350ml dippers in small habitats or 10L buckets in larger habitats, and the mosquitoes were identified based on their morphological characteristics using established taxonomic keys by Gillies and De Meillon and Gillies and Coetzee (Gillies and de Meillon, 1968; Gillies and Coetzee, 1987). Larvae were identified into taxonomic groups of *Anopheles funestus sensu lato* (*sl*), *Anopheles gambiae sl*, and *Culex*, and others. Pictorial data was also collected for all the habitats.

**Figure 3:**
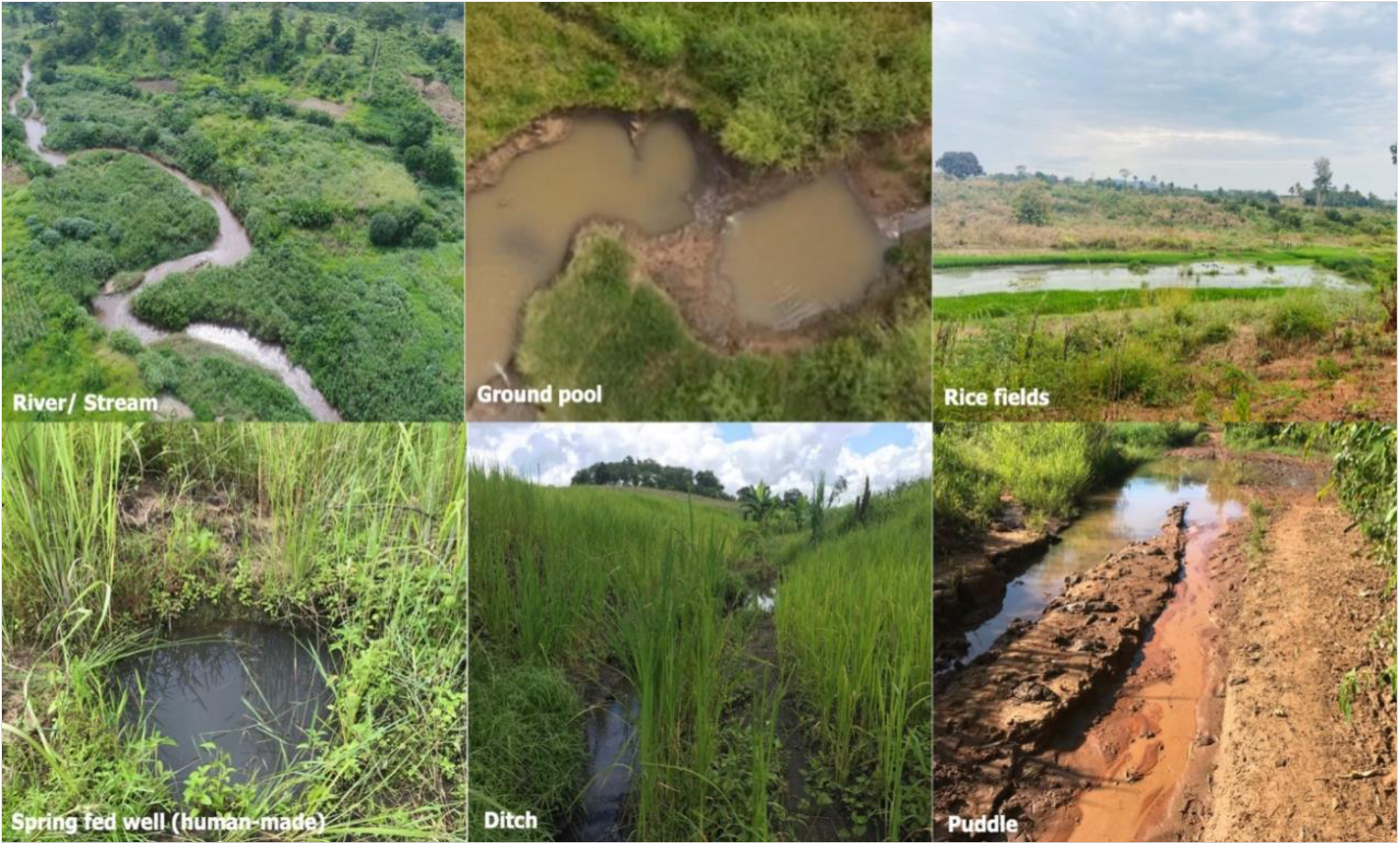
Common types of aquatic habitat found in the study areas.

### Assessment of how communities use the water bodies occupied by *Anopheles* mosquitoes

Once the main habitats occupied by the dominant malaria vectors, *An. funestus* were identified, follow-up observations were done identify and estimate proportion of those habitats that were being used for domestic activities. This was done by directly observing and recording environmental indicators such as footprints, hoof prints, and signs of human and livestock waste within 10m around the habitats, using a prepared checklist. Additionally, we conducted several unstructured interviews with consenting community members living near the habitats to understand how they use them. During these interviews, we gathered information on both the frequency of use, and type of use activities conducted.

Following these initial assessments, we conducted focus group discussions (FGDs) with community members to gain deeper insights into their perceptions of malaria transmission risks within their homes and communities, the connection between local water sources and malaria, and their methods for mitigating transmission risks. Additionally, participants’ attitudes towards the potential and practicality of three LSM approaches were evaluated.

The discussions were structured into three main sections: Initially, participants shared their understanding of malaria transmission, factors contributing to its persistence, and their efforts to mitigate these risks. The second part focused on identifying different sources of mosquito larval habitats and strategies for their control. Finally, participants evaluated the feasibility and effectiveness of the three LSM strategies; larviciding, habitat manipulation (source reduction), and habitat modification (habitat removal) in their community contexts. To foster meaningful dialogue, facilitators provided definitions of each LSM strategy and addressed participant questions before discussions commenced. Participants then shared their perspectives on the appropriateness and potential implementation of these approaches, offering specific recommendations on the contexts and conditions under which each method could be effectively applied.

Altogether, a total of nine FGD sessions were conducted; six with community members (three with males and three with females separately) and one each with local fishermen, pastoralists, and brick-makers. These groups were selected to represent major uses of the water bodies. Each session consisted of eight to ten participants and lasted between one and two hours. Most discussions took place within the participants’ communities (at the village leader’s offices) and discussion with fishermen and brickmakers, participants were invited at Ifakara Health Insistute’s offices. All discussions were audio-recorded for further processing, and detailed notes were taken by at least two facilitators during each session.

### Data processing and analysis

We integrated the analysis of quantitative survey data (done in R statistical software version 4.2.3 (R Core Team, 2019) and qualitative analysis using NVivo software version 12 (*NVivo - Lumivero*, no date). Throughout the analysis process, data weaving and inferencing techniques were employed, integrating information from both components of the study to develop a comprehensive understanding from the viewpoint of the respondents.

For the quantitative data, descriptive statistics were used to summarize the mosquito aquatic habitats and the proportion of those utilized by communities. This included the proportions of all surveyed habitats, those containing mosquito larvae, and specifically, those containing *An. funestus*. The descriptive analysis was extended to categorize the habitats based on their usage by community members for different purposes.

For the qualitative data on the other hand, the audio recordings of the FGDs were transcribed by SK and AS, then reviewed by NFK and FT. During analysis, thematic coding was employed to identify key themes and patterns. Prior to analysis, a code book was developed using both deductive and inductive methods; whereby deductive codes were developed from the objectives of the study and the discussion guide, and inductive codes were developed through a thorough review of the transcripts. Similar codes were subsequently grouped into broader themes and categories that emerged from the data. The coding process was done by NFK and FT. Main themes identified included: i) community understanding about mosquito ovipositing behaviour and their aquatic habitats., ii) participants’ views of the applicability, effectiveness and challenges associated with the three LSM approaches. Direct quotations from the participants were used to support and provide context to the themes

### Ethical considerations

Ethical approval for the study was obtained from the Ifakara Health Institute Institutional Review Board (Ref: IHI/ IRB/No: 26-2020) and the Medical Research Coordinating Committee (MRCC) at the National Institute for Medical Research-NIMR (Ref: NIMR/HQ/R.8a/Vol. IX/3495). Before commencing data collection, permission was obtained from the District Medical Officers (DMO) and subsequently from each village executive officer (VEO). The VEOs assisted in selecting participants for the focus group discussions (FGDs) based on our established criteria. Written informed consent was obtained from all participants prior to their involvement in the FGDs. Additionally, consent for taking pictures of the community members during the observations, surveys and FGDs were obtained.

## Results

### Survey of aquatic habitats in the study areas

The entomological survey identified 931 aquatic habitats, of into six categories, namely: river streams, ground pools, dug pits, rice fields, ditches, and puddles (Figure 3 and Table 2). Nearly three quarters (73%, n = 612) of all the habitats contained mosquito larvae or pupae, and among these 23% (n = 213) contained *An. funestus*.

**Table 1:** Distribution of total habitat surveyed and habitats that had at least mosquito larvae and habitats that had atleast one *An. funestus* mosquitoes.

| Habitat type | Water bodies surveyed | Water identified used by community | Larval habitats | Habitats with *An. funestus* |
| --- | --- | --- | --- | --- |
|  | n (%) | n (%) | n (%) | n (%) |
| River streams | 376 (41) | 353 (42) | 226 (37) | 112 (52) |
| Ground pools | 37 (4) | 33 (4) | 31 (5) | 15 (7) |
| Dug pits | 208 (22) | 188 (22) | 135 (22) | 14 (7) |
| Rice fields | 65 (7) | 64 (8) | 53 (8) | 12 (6) |
| Ditches | 208 (22) | 178 (21) | 139 (23) | 57 (27) |
| Puddles | 37 (4) | 21 (3) | 28 (5) | 3 (1) |
| Totals | 931 (100) | 837 (100) | 612 (100) | 213 (100) |

**Table 2:** Community perspectives on different approaches to larval source management, for malaria vector control.

| Approach | Perceived Benefits | Concerns | Specific recommendations |
| --- | --- | --- | --- |
| **Larviciding** | ▪ Reduces mosquito populations and malaria cases,<br>▪ Targets roots of the problem,<br>▪ Can provide broad area coverage,<br>▪ Balances mosquito control with water needs. | ▪ May have health and environmental safety concerns,<br>▪ May affect aquatic life and livestock,<br>▪ Requires significant cost and labour,<br>▪ Could be limited by community scepticism. | ▪ Educate and engage the community,<br>▪ Involve locals<br>▪ Provide safe use guidelines,<br>▪ Plan and monitor strategically,<br>▪ Ensure effective communication between authorities, communities, and scientists. |
| **Habitat Manipulation** | ▪ Directly controls mosquito breeding sites,<br>▪ Practical and enhances cleanliness,<br>▪ Deters dangerous animals. | ▪ There may be legal and environmental concerns,<br>▪ Is impractical during rainy seasons,<br>▪ Might impact the livelihoods of specific groups, e.g. fishermen. | ▪ Promote community education and participation,<br>▪ Collaborate with government agencies,<br>▪ Establish specific cleanup times and calendar of activity,<br>▪ Adapt methods to seasons and geographical areas, for feasibility. |
| **Source Reduction/**<br>**Habitat Removal** | ▪ Reduces mosquito populations sustainably<br>▪ Leads to a cleaner environment,<br>▪ Decreases malaria cases,<br>▪ Can be implemented in designated areas. | ▪ Is a challenge because the water bodies also serv other purposes,<br>▪ Risks of new breeding sites,<br>▪ There might be community resistance,<br>▪ It is time and labor-intensive. | ▪ Promote education and active community involvement<br>▪ Designate activity areas and provide alternatives<br>▪ Implement regulations and get local leaders to participate<br>▪ Consider and preserve beneficial habitats. |

**Table 3:**
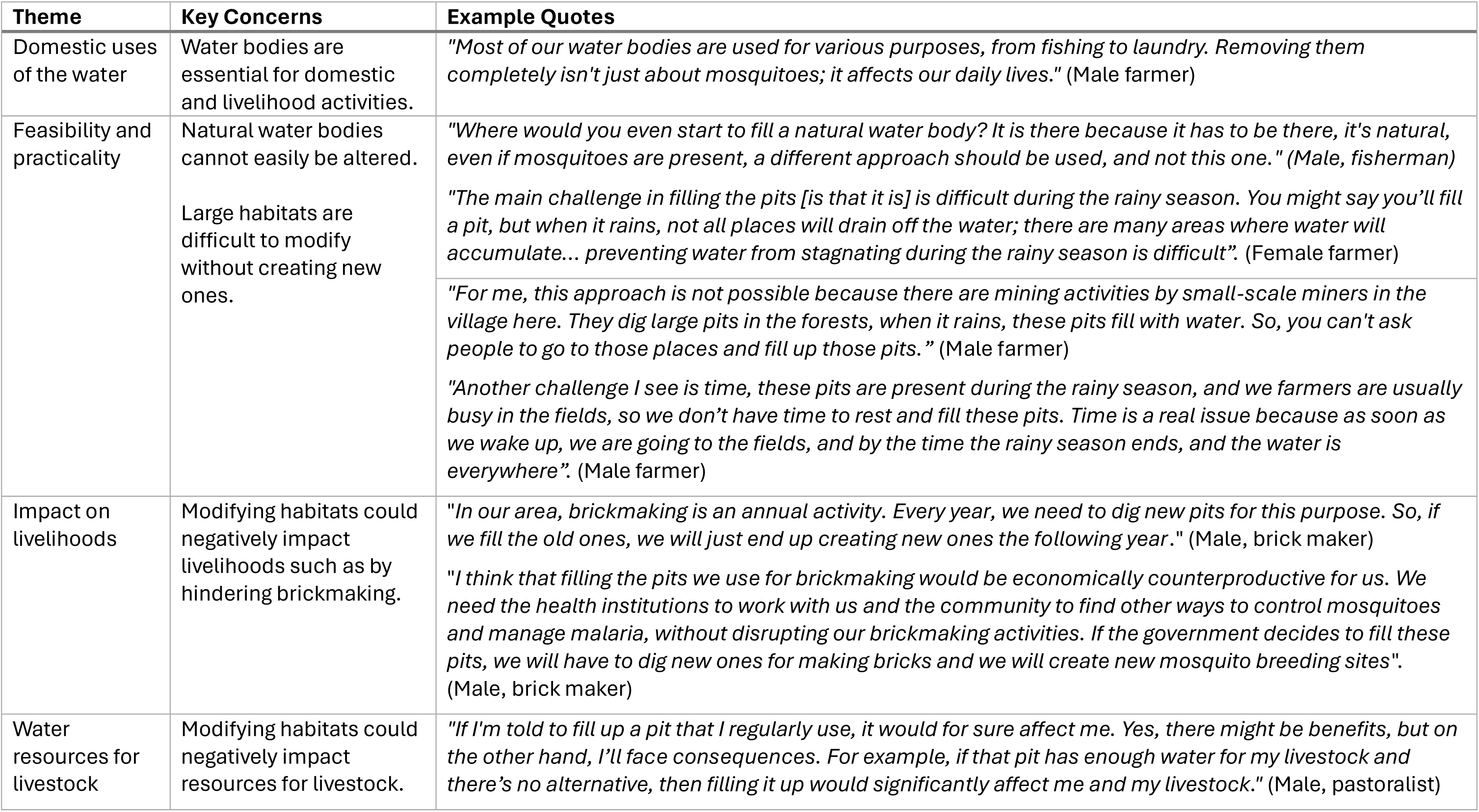
Community concerns regarding source reduction through habitat modification or removal.

In the survey conducted, river streams were identified as the most prevalent aquatic habitats, accounting for 41% (n = 376) of the total, with 37% (n = 226) of these streams serving as larval habitats, and 52% (n = 112) of these larval habitats containing *An. funestus* larvae. Ground pools and ponds represented a smaller fraction, constituting 4% (n = 37) of all identified habitats, with 7% (n = 15) found to be containing *An. funestus* larvae. Human-made pits constituted 22% (n = 208) of aquatic habitats, with 22% of these (n = 135) being larval sites, and 7% (n = 14) of which contained *An. funestus* larvae. On the other hand, rice fields comprised 7% (n = 65) of habitats, with 8% (n = 53) identified as larval sites and 6% (n = 12) harboring *An. funestus* larvae.

Another common habitat type was ditches, which accounted for 22% (n = 208) of all habitats and 23% (n = 139) of all larval habitats. More than half of the mosquito-infested diches ( 57% (n = 78)) were found to have of these containing *An. funestus* larvae. Lastly, puddles formed 4% (n = 37) of habitats, with 5% (n = 28) serving as larval

### Community uses of the different water resources-results of the unstructured interviews and direct observations

The community uses of the different water bodies, which were also found to be containing mosquito larvae are presented in Table 1. The community members used the water from these aquatic habitats for various purposes – of the 931 surveyed, our observations revealed that 90% (n = 837) were being used by community members for one or more purposes (Table 1). Some of the common community uses for the habitats included: source of water for domestic activities such as drinking, cooking, washing dishes and clothes, and bathing, which accounted for 37% (n = 306) of uses; crop irrigation, representing 27% (n = 223); watering livestock, 60% (n = 505); fishing at 37% (n = 311); and brick making at 16% (n = 132) (Figure 4).

**Figure 4:**
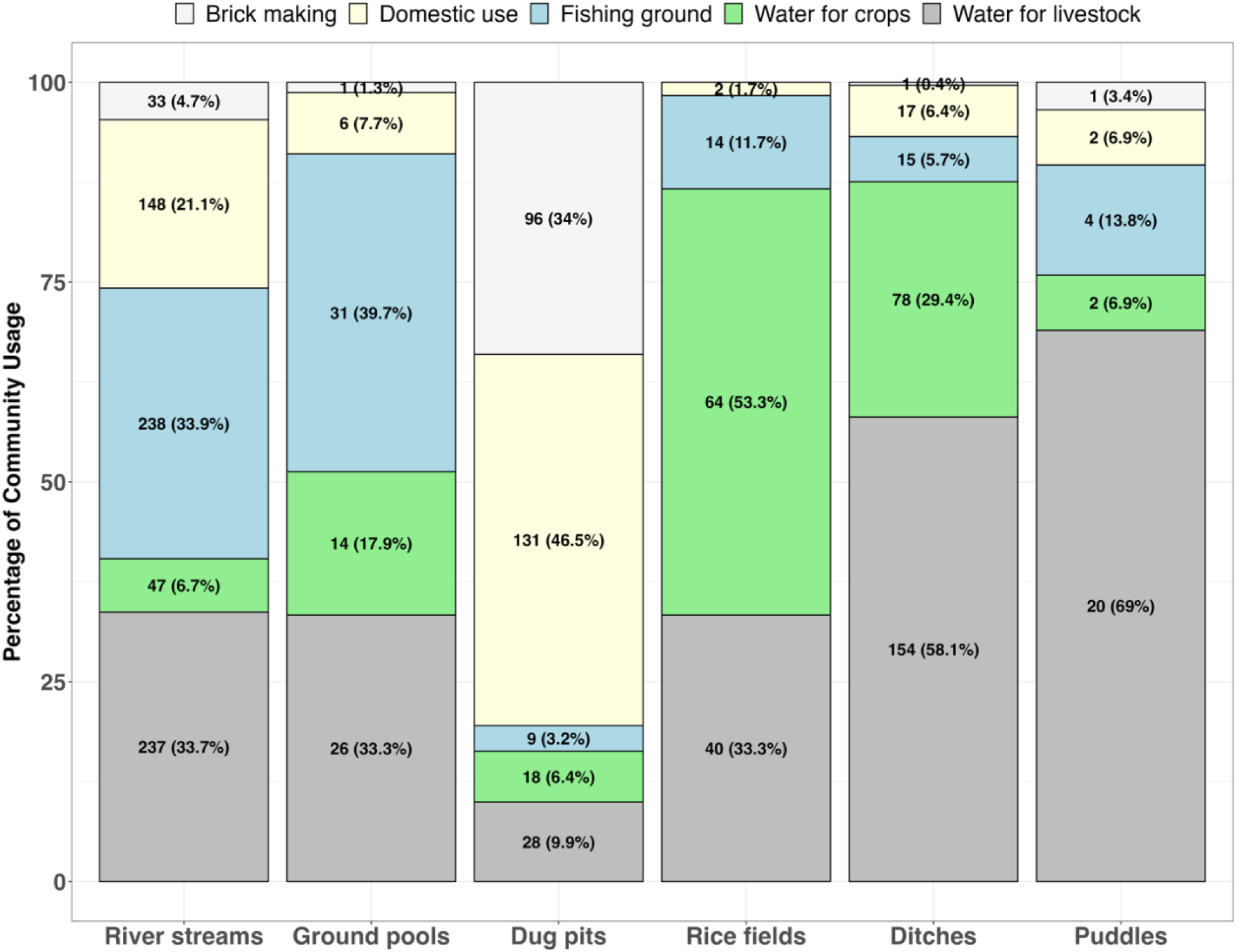
Distribution of different habitat types and those utilized by both *An. funestus* and the community. This figure provides a quantitative summary of the use of different aquatic habitats serving the community for various needs.

**Figure 5:**
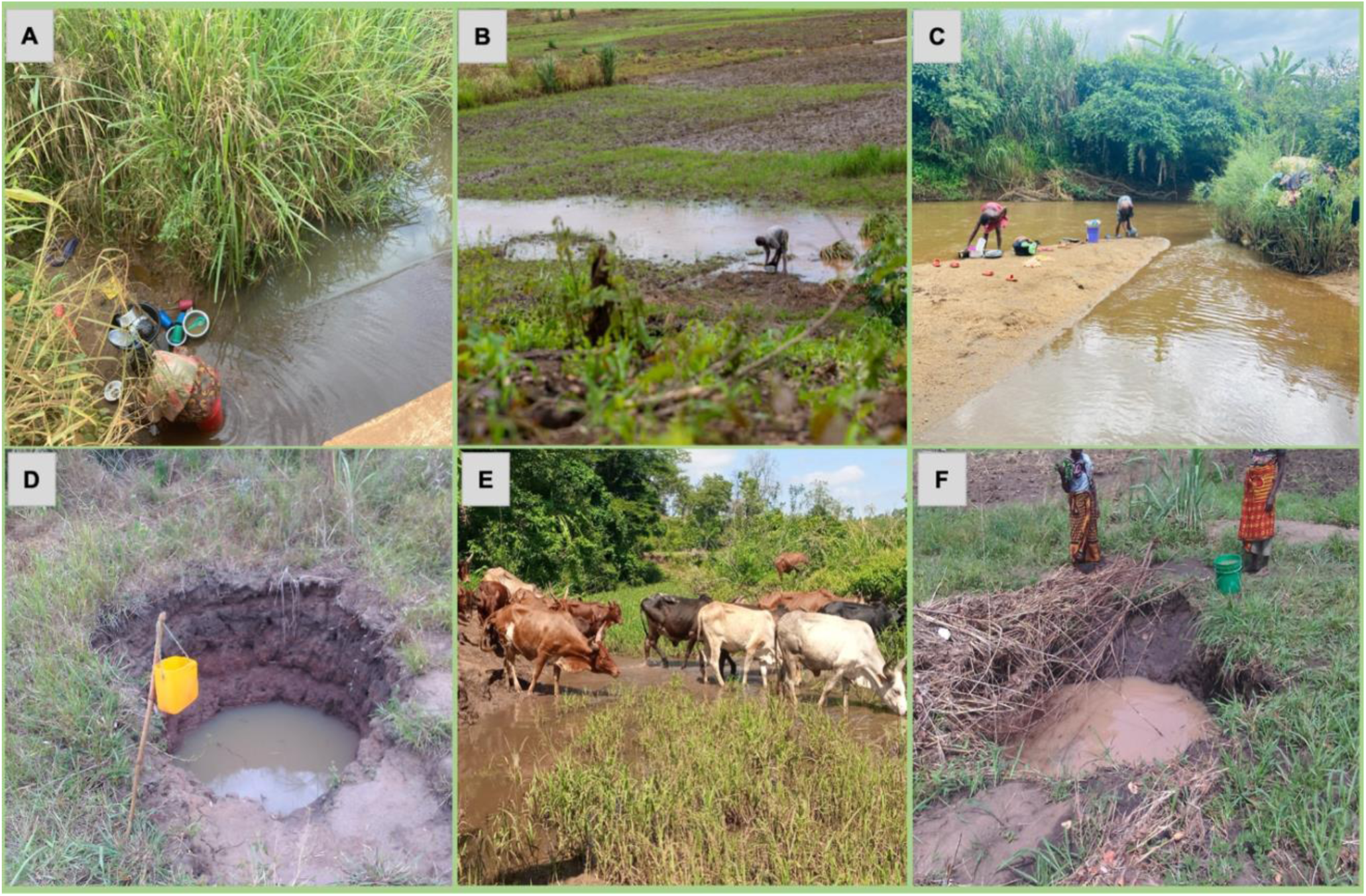
Community dependence on the water bodies identified as also being aquatic habitats. This figure depicts various communal activities conducted in different aquatic environments, illustrating the interplay between daily life and potential mosquito breeding sites. Examples include (A) washing dishes beside river streams, (B) cleaning dishes within flooded rice fields, (C) laundering clothes by riverbanks, (D) fetching drinking water from dug pits, (E) providing water for livestock at river streams, and (F) collecting water from dug pits for household use.

Nearly half of the river streams were used for activities such as fishing, cattle grazing, and domestic needs. The ground pools, including those with *An. funestus* larvae, were commonly used for fishing and cattle grazing. On the other hand, human-made pits served multiple purposes, primarily domestic water uses and brick making.

Approximately 30% (n = 154) of ditches were actively used by community members for cattle grazing and agriculture (Figure 4).

### Results of the Focus Group Discussions regarding community dependance on the different water sources

The focus group discussions comprised a total of 85 participants, consisting of 54 males and 31 females. The age range (determined for 80 of the 85 participants) was 19 to 71 years, with a mean age of 37.92. The majority of respondents had primary education (74%, n=63), while smaller groups had secondary education (19%, n=16) or no formal education (7%, n=6). In terms of marital status, 56% (n=45) were married, followed by 27% (n=23) who were not married, with smaller proportions being divorced (5%, n=4), widowed (7%, n=6), or unidentified (8%, n=7). In terms of occupation, 55% (n=47) were farmers, with others engaged as fishermen (12%, n=10), brick makers (12%, n=10), pastoralists (10.5%, n=9), and various other occupations (10.5%, n=9).The focus group discussions (FGDs) confirmed patterns of community dependence on various water sources, consistent with observations from direct observation and unstructured interviews. During the FGDs, in the community members explained that they relied largely on river streams, dug pits and, in some cases, large pools to obtain water for different purposes including drinking, cooking, watering animals, bathing, and economic activities such as agriculture and construction activities. This dependence on specific water sources was driven by necessity and availability, as one participant voiced the lack of choice in their water source options:

> *“We use what we use because we must, not because it is what we would choose if we had other better and safer options.” (Female farmer)*

The availability and type of water sources varied by location, as revealed by participants in different areas. In some regions, community members utilized groundwater pumps, locally referred to as “*Mdundiko*,” installed by the government, to meet some of their water needs. In contrast, other areas predominantly relied on natural water sources such as rivers, streams and spring-fed wells for most domestic requirements.

Participants noted that a key factor determining the type of water source used was proximity to the village center or main roads. Communities closer to these areas generally accessed more reliable and cleaner water compared to more marginalized ones. One participant described their situation, stressing these disparities:

> *“Our lives in the remote farming area are different from those in the town [here referred to small towns], we in the interior village we dig our wells, and we rely on stream channels, but those in urban areas have pumped water “Mdudindiko” which they use for their domestic needs.” (Female farmer)*

### Community understanding of mosquito reproduction and larval sites

The majority of FGD participants understood that mosquito reproduction involves mating between males and females, followed by females laying eggs in water where they hatch into larvae, developing in aquatic habitats before emerging as adult mosquitoes. Most were familiar with common larval sites such as pits, river streams, and large water bodies. However, some participants lacked such understanding, and sometimes they would mention other unlikely places such as areas with dense vegetation like bushes, pit latrines (that are often not used by malaria vectors), dark and moist places, and corners of the houses. This confusion indicated a mix-up between mosquito breeding sites and the areas where adult mosquitoes are commonly found, as explained by a participant:

> *“From what I know, mosquitoes prefer dark and damp places, particularly those with vegetation. When we clear these areas, the mosquitoes find their preferred habitats disturbed, so they tend to move away, often relocating away from our homes” (Male, farmer)*.

The majority of participants reported observing mosquito larvae in water bodies and associated the presence of larvae with adult mosquitoes. Many respondents noted that they frequently spotted larvae while performing daily chores and were able to identify them as mosquito larvae due to the abundance of adult mosquitoes around water sources, as explained by one of the participants:

> *“I’ve seen mosquito larvae while fetching water from the river. I’ve seen them attached to grasses along the river’s edge, when you disturb the grasses, a group of mosquitoes will fly, these are the same mosquitoes that come to our homes to look for blood.” (Female farmer)*

### Perceptions and recommendations on LSM for malaria control

Discussions on the potential of LSM for malaria control focused on the participants’ views of the applicability, effectiveness, challenges, and recommendations associated with the three LSM approaches. Generally, the participants expressed varying levels of interest for the different LSM approaches (Table 2).

#### Larviciding

Regarding larviciding, the majority of participants expressed enthusiasm, viewing it as a practical option for their communities. They recognized its potential benefits in reducing malaria by linking the use of larvicides to a decrease in mosquito populations. Participants understood that fewer mosquitoes would likely lead to reduced malaria transmission. This proactive approach to lowering malaria risk was widely acknowledged, as said by one of the participants:

> *“Now, there are many puddles and mosquitoes, suppose we apply the larvicide in this area and then we decide to promote its use in other areas too, as we continue to do this, the number of mosquitoes will decrease, and then there will be a reduction in malaria cases.” (Male farmer)*

Many participants believed that addressing the mosquito problem at its source, by preventing larvae from emerging as adults, would be more effective way of controlling malaria compared to ITNs. This viewpoint was commonly relayed in association with the phrase of “*prevention is better than cure*“; highlighting that stopping mosquitoes from maturing into adults, is a preferable strategy. Community members deemed larviciding to be an appropriate approach for targeting mosquito in their habitats, provided that the chemicals used were safe for humans and animals as this participant said:

> *“In my opinion, it is suitable, indeed suitable, to treat mosquito habitats and that is how we could benefit. The mosquito will not be able to emerge, and we will not have malaria, as people say, ‘prevention is better than cure’, it is just like that.” (Female farmer)*

During the discussions, participants expressed concerns about managing larval habitats in remote or forested areas that are often hard to reach and overlooked in community cleaning efforts, such as those organized by local government authorities. This shows the need for strategies that can identify and treat all habitats, including those that are inaccessible. Participants also noted the limitations of localized measures in addressing all larval habitats and emphasized the potential of larviciding to cover more extensive geographic areas.

> *“If we apply the larvicide here, but in the forest, there is another unseen pond! What do we do? The chemical’s efficacy will end, but then the mosquitoes will move and start new habitats there, right? so, the advantage of chemical can be applied in every habitat at the same time.” (Female farmer)*

Participants preferred larviciding partly because they viewed complete removal of mosquito habitats (source reduction) and habitat manipulation as impractical, given the community reliance on these water sources. They appreciated larviciding for its ability to control mosquito populations while maintaining access to essential water resources, highlighting this balance as a significant advantage, as described by one of the participants:

> *“Some of these water habitats we created ourselves because we need them for our daily livelihood, if they keep the mosquito, that was not our intention and so the mosquitoes have to be killed while ensuring the water remains safe and usable for our various purposes.” (Female farmer)*

Despite the general acceptance of larviciding, the focus group discussions (FGDs) revealed some concerns among participants, particularly regarding the use of chemicals to control mosquitoes. First, they questioned whether treated water would still be safe for domestic use and agricultural activities, or their livestock or fish as these participants said:

> *“In our current environment, I don’t think it’s possible because that’s a chemical, but those same water bodies we mentioned are the primary water sources for the community, and then the same water bodies are oviposition sites for the mosquitoes. If the larvicide harms livestock, people might refuse to use it.” (Female, farmer)*.

> *“Because we don’t know if the chemicals, even if they are brought to target mosquito breeding sites, will kill the fish or pose risks to humans, because we lack knowledge and understanding.” (Male fishermen)*.

Secondly, expressed concerns about the feasibility of implementing larviciding in low-income communities due to the required technical expertise and financial resources. In particular, they noted that where multiple larviciding treatments are required, this would be challenging due to lack of continuous financial resources:

> *“The method of applying chemicals is technical and requires financial resources, this method is more realistic, even the larvae of mosquitoes would decrease very fast. However, consistent application is essential, as mosquito populations grow, and the power of the chemical diminishes over time. To prevent mosquitoes from returning, regular reapplication of the chemical is necessary, which needs funds.” (Male farmer)*.

Lastly, the discussions also revealed a general skepticism towards larvicides brought from outside the country, especially fueled as a result of the aftermath of COVID-19, and the skepticism towards COVID vaccine. The participants wondered how communities would be convinced to accept this intervention, as this participant said:

> *“Just like with the coronavirus vaccine, many of us, including myself, were hesitant… How will communities accept these chemicals in the water we drink?” (Male, farmer)*.

### Habitat manipulation

Habitat manipulation for controlling larval habitats received mixed reactions from study participants. Some saw it as a practical measure, particularly getting rid of useless stagnant water near residential areas, while others raised concerns about its feasibility and legality, especially near protected natural water bodies. Supporters of this approach suggested initiatives like clearing tall grass around water sources and homes, noting these actions were practical in their communities and offered additional benefits beyond mosquito control:

> *“We can manage to clear the stagnant water around our homes. It’s something within our power to do, and it helps reduce the mosquito problem in our immediate surroundings.” (Female farmer)*

> *“When you clean and remove the grass, even snakes do not stay, thereby creating a safer environment.” (Female farmer)*

The primary concern regarding habitat manipulation, as voiced by most participants, centered on the inaccessibility of certain water bodies, especially those in government-protected areas or regions with land development restrictions. Participants note that there were legal prohibitions against altering vegetation within 60 meters of a river stream to avoid ecological disturbances, emphasizing the regulatory challenges associated with this approach:

> *“We have a nearby river Luli. It has reeds and dense vegetation. Now, that vegetation hosts numerous organisms like snakes, chameleons, and lizards*.

Tanzania National Parks Authority (TANAPA) *cannot allow you to clear vegetation 60 meters around the river streams because it will chase away these animals from their natural habitats*.” (Male farmer)

Moreover, habitat manipulation was also considered impractical during the rainy season when there was often flooding and water everywhere, making it impossible to clear the water or to keep up with vegetation growth as this participant elaborated:

*“During the rainy season vegetation grows fast around water bodies, if you clear it today, it quickly grows back in just days. (Female farmer)*

This approach was especially opposed by fishermen who feared that it could disrupt breeding habitats for the fish. The fishermen explained that vegetation alongside riverbanks provided calm waters and safe havens for fish to lay their eggs. Disrupting this, therefore, could interfere with their livelihoods as this fisherman explained:

> *“Fish prefer to lay their eggs in parts of the river that are calm and have plenty of vegetation. You won’t find fish eggs in fast-flowing waters. If you tell me to clear the grass along the riverbanks today, I will also be disturbing breeding habitats for the fish. This will likely reduce fish reproduction and ultimately harm our income.” (Male fishermen)*

One particular exception was that the pastoralists who participated in these discussions expressed their support for this approach as they deemed it would not have negative impact on their livestock. Reducing vegetation alongside water sources was also perceived as beneficial as it opened up the water for their livestock. However, they too were concerned about whether or not they would have time to do such work, given their nomadic nature and busy schedules as this participant said:

> *“I wonder when we, as pastoralists, would find the time for this task. Every day we’re up early to go grazing and don’t return until evening. If the government decides to undertake this exercise, we will agree, because our animals only go to these places for water.“*

### Source reduction through habitat modification or removal

This approach was the least favored due to multiple reasons: i) most water bodies are utilized for domestic or livelihood purposes, ii) concerns that filling these water bodies would require creating other pits to obtain sand, which could potentially become new larval habitats, and iii) the impracticality of altering natural water bodies (Table 2). Participants noted that it might be feasible for smaller, unused breeding sites like puddles within the community. The potential for habitat modification to completely eliminate water sources was a significant concern, especially given the multifunctional nature of these resources in the community. Consequently, participants deemed this approach both impractical and inapplicable.

Another concern raised regarding habitat modification was that some habitats are too large to modify or remove without creating new potential breeding sites. Other than the river streams and large ponds, other examples of these were the pit holes resulting from brickmaking and mining activities; study participants explained that it would be impossible to find landfills to cover these without creating more pits in the process as this participant elaborated. Participants noted the impracticality of finding adequate landfill materials to fill these large pits without the need to excavate additional areas. Additionally, the approach was deemed unsuitable during the rainy season due to frequent flooding that enlarges water bodies, complicating any efforts to control habitats.

Furthermore, participants emphasized the practical challenges and the effort required to engage in habitat modification amidst their busy agricultural schedules. Brickmakers, in particular, opposed this strategy because it threatened their livelihood. They rely on pits filled with water for brickmaking and create new pits annually, suggesting that filling these would directly impact their income. Similarly, pastoralists expressed concerns about the adverse effects on their livestock, emphasizing their reliance on these water sources and preferring to maintain them for animal use.

### Broad recommendations by community members regarding LSM

Key recommendations made by participants during this study are summarized in Table 4. To ensure effectiveness of LSM strategies in the study communities, the participants members emphasized the importance of raising awareness about the techniques for malaria control. They suggested awareness campaigns, e.g. through community meetings, to address potential impacts that LSM might have on people, livestock, and the environment. Participants also advocated for clear understandable guidelines on the use of larvicides, including their frequency and safe application timing, as relevant to specific settings. Thirdly, they emphasized the importance of involving locals in program implementation to foster trust and ownership, noting that community support would increase if implementation were led by familiar faces.

**Table 4:** Recommendations from community members regarding larval source management.

| Theme | Recommendations | Example Quotes |
| --- | --- | --- |
| **Awareness and Education** | Conduct awareness campaigns to educate and answer questions about the impacts of LSM on people, livestock, and the environment. | *"Community members might reject [the idea] initially due to a lack of understanding. Education should be provided first, then once people understand, they will accept it."* (Female farmer) |
| **Guidelines for Larviciding** | Provide clear guidelines on the dosage and timing of larvicides to ensure safe usage. | *"We need guidance to inform us on the appropriate time for spraying and the correct dosage to avoid unintentional harm to other beings."* (Male farmer) |
| **Local Involvement** | Train and involve local community members in LSM implementation to build trust and ensure program ownership. | *"If I see a local person, someone from our own village, doing the larviciding, I will have more faith than if it were strangers who I don't know their intentions."* (Male farmer) |
| **Timing of Implementation** | Carefully plan the timing of LSM to align with seasonal variations and community readiness. | *"The rainy season is the best time for spraying chemical, as many puddles and mosquito breeding places are at its peak. That's when the larvicide should be applied."* (Female farmer) |
| **Government Mandate** | Advocate for government-mandated LSM activities, including environmental cleaning and removal of stagnant water. | *"In my view, for people to engage in this exercise, the government should set a specific day for cleaning, like the 'Magufuli Saturday', where everyone knows they should be cleaning around their premises."* (Male farmer) |
| **Alternative Water Sources** | If water sources are modified or removed, provide alternative sources for community use. | *"Abandoned pits near homes can be filled, but those used for community activities, like domestic purposes, watering animals, construction activities like building clinics or schools, can be difficult for the community to agree."* (Female farmer) |

There was a consensus on the need for careful planning of the deployment and timing of LSM to align with seasonal variations and readiness. Participants suggested that different LSM strategies might have different calendars of activities. For instance, while larviciding may be desirable during the rainy season when water sources are plentiful and vector populations are highest, habitat manipulation would be more feasible in the dry season when water bodies are fewer and also reduced in size. To enhance the impact, participants suggested that local governments should mandate regular LSM activities, such as environmental clean-ups – for example, they suggested bi-weekly community cleaning days to encourage broad participation.

Additionally, if water sources are modified or removed, there was a strong recommendation for the government agencies to provide alternative sources and to reallocate activities like brickmaking to minimize environmental risks. The community emphasized that habitat modification should be limited to unused water sources to avoid disrupting local needs. This holistic approach highlights the community’s concern for careful planning and local involvement in LSM initiatives.

## Discussion

Effective larval source management (LSM) necessitates a nuanced understanding and targeted approach to water bodies that are essential for mosquito larval development. However since local communities often depend on the same water bodies for various other purposes, targeting these habitats for vector control requires careful considerations of local societal practices and expectations. The overall goal of this project was to explore how local communities in rural southeastern Tanzania use these water bodies and how this might influence LSM strategies. We focused primarily on habitats frequented by *An. funestus*, first because this is the primary malaria vector in rural south-eastern Tanzania, and second, because the vector species prefers a set of unique habitats that often remain as the sole water supplies during the dry season. Overall, our results illustrate a dual challenge: the critical need for water resources for various community purposes on one hand, and the simultaneous need to effectively manage these resources effectively to limit mosquito breeding.

It was observed that a vast majority of aquatic habitats used by malaria vectors, notably river streams, ground pools, dug pits, rice fields, ditches, and puddles, are integral to the daily lives of local communities; where they are used for washing, fishing, cattle grazing, and even as sources of drinking water. This linking of community life with potential mosquito vector larval habitats underlines the importance of engaging with communities to tailor LSM approaches that will respect their reliance on these habitats (van den Berg *et al*., 2018). While the community members acknowledged the need for effective malaria vector control measures, there was a clear call for these measures to be applied thoughtfully considering the multiple uses of aquatic habitats by local communities. The need for more information about the safety and impact of LSM approaches on daily life was emphasized, pointing towards a gap in communication and education regarding LSM strategies.

The survey revealed that aquatic habitats, such as river streams, ground pools, and human-made pits, play crucial multifunctional roles in community life, serving as essential resources for domestic and agricultural activities as well as breeding grounds for *An. funestus* larvae. This complexity presents significant challenges for malaria control efforts, such as habitat manipulation or larviciding, which must balance ecological impacts with community needs. Community interactions with these water sources varied widely, including uses for drinking, irrigation, brick making, and livestock watering. Notably, river streams and ground pools were frequently used for washing dishes and clothes, reflecting their accessibility and utility, which aligns with findings from Kenya highlighting similar dependencies on aquatic habitats for daily chores (Imbahale *et al*., 2010). Dug pits and ditches were commonly associated with brick making and agriculture, indicating their importance in economic and food production activities. This diverse use cases underline the significance of water bodies to community livelihood and underscores the need for LSM strategies that effectively control malaria vectors while being culturally and practically acceptable to the communities they serve (Walker and Lynch, 2007; Fillinger *et al*., 2009; Fillinger and Lindsay, 2011).

Building on the varied use of aquatic habitats, it was noted that community members had a solid understanding of the mosquito lifecycle including ability to distinguish between aquatic stages and adult life stages. They could identify mosquito larvae and understood the direct relationship between the presence of larvae and the subsequent increase in adult mosquito populations. This level of awareness is supported by findings from other research, which has consistently shown a considerable understanding of malaria transmission dynamics within malaria endemic communities (Imbahale *et al*., 2010). For example, research conducted in similar settings have reported that, local communities are often aware of mosquito breeding sites and their link to the risk of malaria (Finda *et al*., 2019; Tarimo *et al*., 2023). However, these studies also indicate variation in the depth of knowledge and its application towards preventive practices; suggesting that while awareness is widespread, its effective interpretation to reduce malaria risk may differ from one community to another.

The study underscored significant challenges in LSM strategies, particularly habitat manipulation and source reduction, due to their potential to disrupt community livelihoods. Modifying water bodies used for brick making and livestock could adversely affect local economies and animal welfare, emphasizing the need for a careful balance between effective vector control and community sustainability. Larviciding, although favored for its perceived straightforwardness and effectiveness in controlling mosquito populations, raised concerns about the safety of water post-treatment, impacting livestock and aquatic life. These concerns about the safety of larvicides and their potential impact on human, animal, and environmental health, including aquatic life, echo broader challenges previously documented (Gowelo *et al*., 2020; Hakizimana *et al*., 2022). However, studies in regions with similar living conditions indicate that people generally accept the use of larvicides, provided they do not negatively impact the environment or their way of life (Berlin Rubin *et al*., 2020; Gowelo *et al*., 2023).

Nonetheless, the persistence of environmental and health safety concerns, which have also been observed in other studies highlights the importance of community education and involvement in LSM to ensure acceptance and understanding of these methods (Geissbühler *et al*., 2009; Berlin Rubin *et al*., 2020; Hakizimana *et al*., 2022; Gowelo *et al*., 2023).

Habitat manipulation, recognized under initiatives like the “*Jumamosi ya Magufuli*” campaign in Tanzania, an initiative started by Tanzania former President, which promotes environmental cleanliness, was also seen as a viable LSM approach (Mvomero, 2023). This highlights how existing policy and government information campaigns can be harnessed to promote LSM based on habitat management.

However, its application is limited near natural water sources that are legally protected, stressing the need to consider environmental regulations in LSM planning (Walker and Lynch, 2007). Moreover, concerns from some sections of the communities, for example fishermen who were concerned about the potential negative impact on fish breeding habitats, illustrate additional complexities in applying LSM approaches to river streams, which comprise a significant water source in rural communities. These findings suggest that while LSM strategies are essential, they must be adaptable and sensitive to both ecological and community contexts.

While source reduction approaches are considered the most effective strategy for mosquito control because it completely removes larval habitats (Killeen, Seyoum and Knols, 2004; Tusting *et al*., 2013), this study suggests it was also the least preferred method among community members. The main concern was its potential impact on livelihoods and daily activities. Nearly all identified larval habitats were also used by community members for different purposes. Farmers rely on these water sources for both irrigation and domestic purposes; pastoralists need them for their livestock; brick makers use them in their brick-making processes, and fishermen depend on them for their catch. Similar patterns were observed in Malawi, where community dependence on mosquito larval habitats for various activities was reported (Gowelo *et al*., 2020). This highlights the need for a careful balance between implementing public health measures to combat malaria and ensuring the well-being of communities, especially in areas where livelihood and daily activities are connected to the environment (Walker and Lynch, 2007; Koenraadt, 2021).

Community members emphasized the importance of raising awareness and providing education about the potential risks and benefits of all LSM approaches. They advocated for open communication and active community engagement in mosquito control efforts to ensure broad understanding and involvement in the implementation process (Walker and Lynch, 2007). Additionally, they pointed out the need of carefully scheduling these activities, selecting time periods for implementation both when the intervention will be more effective and when the majority of the community can actively participate. This highlights the necessity of adapting interventions to the dynamic nature of mosquito larval habitats and human activities This concurs with the wider body of evidence indicating that vector control initiatives are more successful and relevant to local needs when the community is well-informed and directly involved in the mosquito control efforts (van den Berg *et al*., 2018). One challenge that could rise with regard to this is balancing the timing of LSM implementation.

Most importantly, community members voiced that if habitat manipulation or modification-based LSM would be pursued as part of mosquito control efforts, the government should ensure the provision of alternative water sources. This demand highlights the communities’ concern over the potential negative impacts such interventions could have on their daily lives, stressing the importance of mitigating these effects through thoughtful planning and the establishment of support systems. It is increasingly recognized that environmental management for malaria control must be integrated with local development needs (Berlin Rubin *et al*., 2020; Koenraadt, 2021; Hakizimana *et al*., 2022). Our findings add to this by proving the importance of not only addressing the public health aspects of malaria control but considering the broader implications on community access to water, agricultural practices, and overall economic well-being. Successful LSM interventions require a holistic understanding of local ecosystems and socio-economic dynamics; ensuring that efforts to combat malaria do not inadvertently compromise the resources upon which communities depend.

Therefore, a trade-off between mosquito control measures and community use of water sources can be significantly mitigated through better investment in water infrastructure. Improving water infrastructure could create a “win-win” scenario by simultaneously addressing malaria control and enhancing other areas of health, such as Water, Sanitation, and Hygiene (WASH), while supporting economic livelihoods (World Health Organization, 2023). By providing reliable and safe alternative water sources, communities would be less dependent on natural habitats that serve as mosquito breeding grounds, allowing for more effective LSM strategies without compromising community needs (van den Berg *et al*., 2018; Gowelo *et al*., 2020).

Moreover, better water infrastructure can improve overall public health by reducing waterborne diseases and providing essential resources for agriculture and livestock, thus boosting local economies (Batterman *et al*., 2009). Integrating LSM efforts with broader development initiatives focused on enhancing water infrastructure would not only facilitate sustainable malaria control but also promote long-term community well-being and resilience (Whittaker and Smith, 2015; Hakizimana *et al*., 2022).

While this study has been the first to extensively explore the interaction between mosquito aquatic habitats and community needs in south-eastern Tanzania, it has some limitations in the methodological approach. The study primarily collected data through direct observations and FGDs with communities from selected villages, purposely chosen for their observable use of water sources that also serve as mosquito larval habitats. This approach was taken to facilitate understanding of the importance of aquatic habitats for human activities would impact the acceptance of mosquito control measures. However, by focusing on these specific settings, the study may have overlooked areas where such habitats play a lesser role in the community’s daily life. Future research should therefore include a more diverse locations, especially those where reliance on mosquito larval habitats for water is not a significant aspect for daily living.

## Conclusions

This study provides valuable insights into community perspectives on LSM for malaria control and elimination efforts. Additionally, it shows the complexities that might arise during the planning and implementation of LSM given the dual role of aquatic habitats as both important community resources and breeding sites for malaria vectors. In settings such as south-eastern Tanzania, where the dominant malaria vector, *An. funestus,* primarily breeds in permanent and semi-permanent habitats such as river streams, large pond and spring-fed pools, our study reveals a clear preference for strategies like larviciding and habitat manipulation, which can more easily be aligned to daily activities and have minimal disruption to local livelihoods. These findings emphasize the importance of community engagement and the need for LSM strategies to be both culturally and environmentally sensitive to achieve community acceptance and sustainability. Furthermore, findings emphasize the need for balanced approaches that respects community practices and environmental considerations. Indeed, engaging communities in the design and implementation of LSM, along with providing education on the safety and efficacy of such interventions, is vital to ensure these strategies do not negatively impact local water resources. Finally, it is important to consider the socio-economic and regulatory constraints, especially regarding protected natural water sources. This calls for adaptable, community-informed strategies that maximize public health benefits while preserving community well-being and environmental integrity. Ultimately, vector control approaches should be designed in a holistic manner, ensuring to integrate the needs, perspectives, and daily lives of the communities it aims to protect.

## Data Availability

Upon reasonable request

## Acknowledgements

We express our gratitude to the community leaders and residents in all the study villages for granting us permission to carry out our work in their localities. We thank Ramadhani Bofu, Christina Makungu, and Bosco Chinkonda, for advising the qualitative aspects of this work. We also extend our thanks to the technicians and volunteers who actively contributed to the successful execution of this comprehensive survey.

## Authors’ contributions

Conceptualization of study and design the research framework: FOO, NFK, FT, and MF. Leading in producing the initial and subsequent drafts: NFK, and FT. Data collection: NFK, KK, FT, LF, SK, and WM. Audio transcription: SK, AS, reviewing the transcripts NFK and FT. Data analysis and interpretation of results: NFK, FT, FOO, and MF. Critical reviewed the manuscript drafts: MF, FOO, HMF, FB, SM, BJM. Supervision throughout the study period: HMF, FOO, FB, MF. All authors read and approved the final manuscript.

## Availability of data and material

Upon reasonable request

